# Intermittent Theta Burst Stimulation (iTBS) and Resting State Functional Connectivity in People Living with HIV/AIDS (PLWHA) Who Smoke Tobacco Cigarettes

**DOI:** 10.1101/2023.05.08.23289662

**Authors:** Gopalkumar Rakesh, Thomas G. Adams, Rajendra A. Morey, Joseph L. Alcorn, Rebika Khanal, Amanda E. Su, Seth S. Himelhoch, Craig R. Rush

**Author notes:** Corresponding Author Gopalkumar Rakesh MD Assistant Professor; Department of Psychiatry 245 Fountain Court; Lexington, KY, 40509; Email –. Disclosures/Conflicts of Interest: None.

## Abstract

**Background:** People living with HIV (PLWHA) smoke at three times the rate of the general population and respond poorly to cessation strategies. Previous studies examined transcranial magnetic stimulation (TMS) over left dorsolateral prefrontal cortex (L. dlPFC) to reduce craving, but no studies have explored TMS among PLWHA who smoke. The current pilot study compared the effects of active and sham intermittent theta-burst stimulation (iTBS) on resting state functional connectivity (rsFC), cigarette cue attentional bias, and cigarette craving in PLWHA who smoke.

**Methods:** Eight PLWHA were recruited (single-blind, within-subject design) to receive one session of iTBS (n=8) over the L. dlPFC using neuronavigation and, four weeks later, sham iTBS (n=5). Cigarette craving and attentional bias assessments were completed before and after both iTBS and sham iTBS. rsFC was assessed before iTBS (baseline) and after iTBS and sham iTBS.

**Results:** Compared to sham iTBS, iTBS enhanced rsFC between the L. dlPFC and bilateral medial prefrontal cortex and pons. iTBS also enhanced rsFC between the right insula and right occipital cortex compared to sham iTBS. iTBS also decreased cigarette craving and cigarette cue attentional bias.

**Conclusion:** iTBS could potentially offer a therapeutic option for smoking cessation in PLWHA.

## Introduction

People living with HIV/AIDS (PLWHA) smoke at three times the rate of the general population^1^. These elevated smoking rates are associated with greater morbidity and mortality^2^. Smoking impacts the progression and outcome of HIV disease and has been identified as the leading contributor to premature mortality among PLWHA^3^. Smoking cessation may be the single most important behavioral health change for PLWHA who smoke^4,5^.

Pharmacotherapy is a pillar of smoking cessation treatment^6^. Evidence regarding pharmacological smoking cessation strategies such as nicotine replacement therapy, bupropion, and varenicline in PLWHA is mixed and limited ^7,8^. Although nicotine replacement therapy has been shown to be effective compared to placebo in people without HIV for smoking cessation for up to six months^9^, controlled trials comparing nicotine replacement therapy to placebo are lacking in PLWHA who smoke. There have been no RCTs (Randomized Clinical Trial) examining efficacy of bupropion for smoking cessation in PLWHA who smoke. Additionally, bupropion’s pharmacokinetic interactions with antiretroviral therapy pose a barrier in PLWHA who smoke^10^. While more effective than placebo, quit rates observed with varenicline remain troublingly low in PLWHA who smoke ^11–14^. Elevated rate of smoking in PLWHA, suboptimal treatment response rates, and lack of adherence to cessation strategies ^15^ underscore a significant need for novel or adjunct interventions for smoking cessation in PLWHA who smoke.

Multiple studies have demonstrated efficacy of transcranial magnetic stimulation (TMS) in decreasing craving for cigarettes and for smoking cessation^16^. Fourteen studies have tested the effects of TMS in people who smoke^17–30^. The majority (12) applied excitatory high frequency TMS ranging from 10-20 Hz ^17–28^. The number of sessions in these studies ranged from 1-18 and cortical targets were heterogenous. Nine studies targeted the left dorsolateral prefrontal cortex (L. dlPFC), one study targeted the left superior frontal gyrus, and two studies targeting the right dlPFC. Moreover, two deep TMS studies targeted the bilateral dLPFC and insula using H coil. All studies, save for one^30^, reported reductions in clinically relevant smoking metrics (e.g., subjective craving, number of cigarettes smoked, abstinence using exhaled CO or cotinine). All nine studies targeting the L. dlPFC showed reduction in clinically relevant smoking metrics ^17–19,21–26^.

Theta burst stimulation (TBS) was approved by the Food and Drug Administration (FDA) for treatment of major depressive disorder (MDD)^31^. Intermittent theta burst stimulation (iTBS) has been shown to be excitatory and 1800 pulses of iTBS has been seminal to a novel accelerated treatment protocol for MDD (Major Depressive Disorder) called Stanford Neuromodulation Treatment^32,33^. To the best of our knowledge, there has been only a single clinical study that utilized iTBS for smoking cessation^29^, which found that four sessions of iTBS (600 pulses) with cognitive-behavioral therapy (CBT) was associated with a significantly greater reduction in smoking urges when compared to sham iTBS with CBT^29^.

Previous studies have shown resting state functional connectivity (rsFC) changes following iTBS between the target site and other brain regions^34–40^. Three studies have shown rsFC changes when iTBS was applied over L. dlPFC^41–43^. The application of three trains of iTBS (600 pulses at 80% resting motor threshold [RMT]) at five-minute intervals reduced L. dlPFC to right anterior insula rsFC when compared to sham iTBS in healthy controls (*n*=28)^41^. Another study applied a single train of iTBS (600 pulses at 90-120% RMT) over L. dlPFC to 18 healthy controls and found that rsFC increased (from baseline) between the L. dlPFC and bilateral caudate^42^. iTBS in ten healthy volunteers (600 pulses at 80 % RMT) over the L. dlPFC revealed increased rsFC between the bilateral superior frontal gyri, and bilateral, middle frontal, inferior frontal, and orbitofrontal gyri immediately after iTBS, compared to rsFC before iTBS^43^.

Although neuroimaging correlates of iTBS in people who smoke cigarettes have not been explored, two published studies investigated neural correlates of 10Hz TMS in people who smoke cigarettes ^44,45^. One study delivered one session of 10Hz TMS (3000 pulses) over the L. dlPFC in eleven people who smoked cigarettes and showed reductions in blood oxygen level dependent (BOLD) activity during a cigarette cue reactivity task in the left nucleus accumbens and right medial orbitofrontal cortex (OFC) when compared to sham TMS ^45^. The other study compared one session of 10Hz TMS versus sham TMS over the L. dlPFC in ten participants who smoked cigarettes and showed decreased resting fractional amplitude of low frequency fluctuation (fALFF) in the right insula^44^, which suggests reduced activity in this region.

Cigarette cue attentional bias (AB) offers a behavioral paradigm to monitor TMS effects in smokers ^46,47^. This bias (AB) is quantified using fixation time on cigarette and neutral cues, via an eye tracker. It can predict the severity of cigarette smoking as well as the chances of relapse^48–50^. Two previous TMS studies comparing 3600 pulses of continuous theta burst stimulation (cTBS) and sham cTBS over the left medial prefrontal cortex (MPFC) when viewing alcohol and cocaine cues, showed decreased seed based functional connectivity (MPFC as seed) with cTBS compared to sham cTBS ^51,52^. In recent years, fMRI studies have also shown the right insula to be a important for interoceptive awareness, cigarette cue reactivity and craving in participants who smoke ^53–55^.

To the best of our knowledge, no published studies have examined the effects of iTBS on rsFC in subjects who smoke cigarettes, no studies have examined the effects of iTBS of the L. dlPFC on cigarette cue attentional bias, and no studies have evaluated the effects of TMS or iTBS among PLWHA who smoke. The following pilot study aimed to examine within subject effects of a single session of iTBS (1800 pulses at 120% RMT) administered to L. dlPFC using individualized neuronavigation on rsFC, cigarette cue attentional bias, and cigarette craving. In line with previous studies, our study hypotheses were as follows: 1) iTBS would alter rsFC between L. dlPFC and the right insula when compared to sham iTBS; 2) iTBS would alter rsFC between right insula and the dorsal cingulate cortex when compared to sham iTBS; 3) iTBS would decrease fixation time on cigarette cues and cigarette cue AB when compared to sham iTBS, and; 4) iTBS would decrease craving for cigarettes when compared to sham iTBS.

## Methods

### Participants

Eight participants were recruited from the Bluegrass Care Clinic within the University of Kentucky Medical Center. The average age of participants (comprising seven males and one female) was 42.88 years. All five participants who returned to receive sham iTBS were male. Average Fagerstrom Test for Nicotine Dependence (FTND) score was 5.75 for the sample and participants endorsed smoking an average of 22.75 cigarettes daily. See **Table 1** for details.

**Table 1.** Demographics.

|  |  |
| --- | --- |
| Age in years, mean (*SEM* *) | 42.88 (3.45) |
| Number of males : number of females | 7:1 |
| Years of education, mean (*SEM*) | 14(1) |
| Right Handedness | 8 |
| Fagerstrom Test for Nicotine Dependence (FTND), mean (*SEM*) | 5.75 (0.96) |

\**SEM* – Standard Error of the Mean

### Experimental Protocol

The study protocol was approved by Medical Institutional Review Board (IRB) at University of Kentucky (IRB#64473) and was registered on clinical trials.gov (NCT04936594). Participants were recruited from the Bluegrass Care Clinic affiliated with University of Kentucky Medical Center. See Supplementary Material Section 1 for details regarding the number of patients who screened eligible for the study or dropped out. For a list of inclusion/ exclusion criteria, please see Supplementary Material Section 2. **Figure 1** summarizes the three-day experimental protocol. After completing a phone screen to determine eligibility, participants were scheduled for Day 1 procedures, which consisted of informed consent, collection of demographic details, measuring carbon monoxide (CO) using a Smokerlyzer Breath CO Monitor (Bedfont Scientific Ltd., Rochester, England), measuring blood alcohol level (BAL) using a Breathalyzer, administering Fagerstrom Test for Nicotine Dependence (FTND) and an attentional bias screen (AB Screen). All study participants were scheduled to receive iTBS a week from Day 1 (Day 2) and sham iTBS five weeks from Day 1 (Day 3). iTBS and sham iTBS sessions were separated by four weeks to limit carryover effects. Day 2 consisted of 1) resting state fMRI brain scans before and after iTBS (rs-fMRI Time 1 and rs-fMRI Time 2 respectively), 2) attentional bias before and after iTBS (AB Time 1 and AB Time 2 respectively) and 3) cigarette craving assessment using the short form version of tobacco craving questionnaire before and after iTBS (TCQ-SF Time 1 and TCQ-SF Time 2 respectively). Day 3 was procedurally the same as Day 2 except sham iTBS was administered instead of iTBS and rs-fMRI scan were only completed after sham iTBS (rs-fMRI Time 3). All participants received iTBS first and sham iTBS subsequently but were blinded regarding treatment.

**Figure 1.**
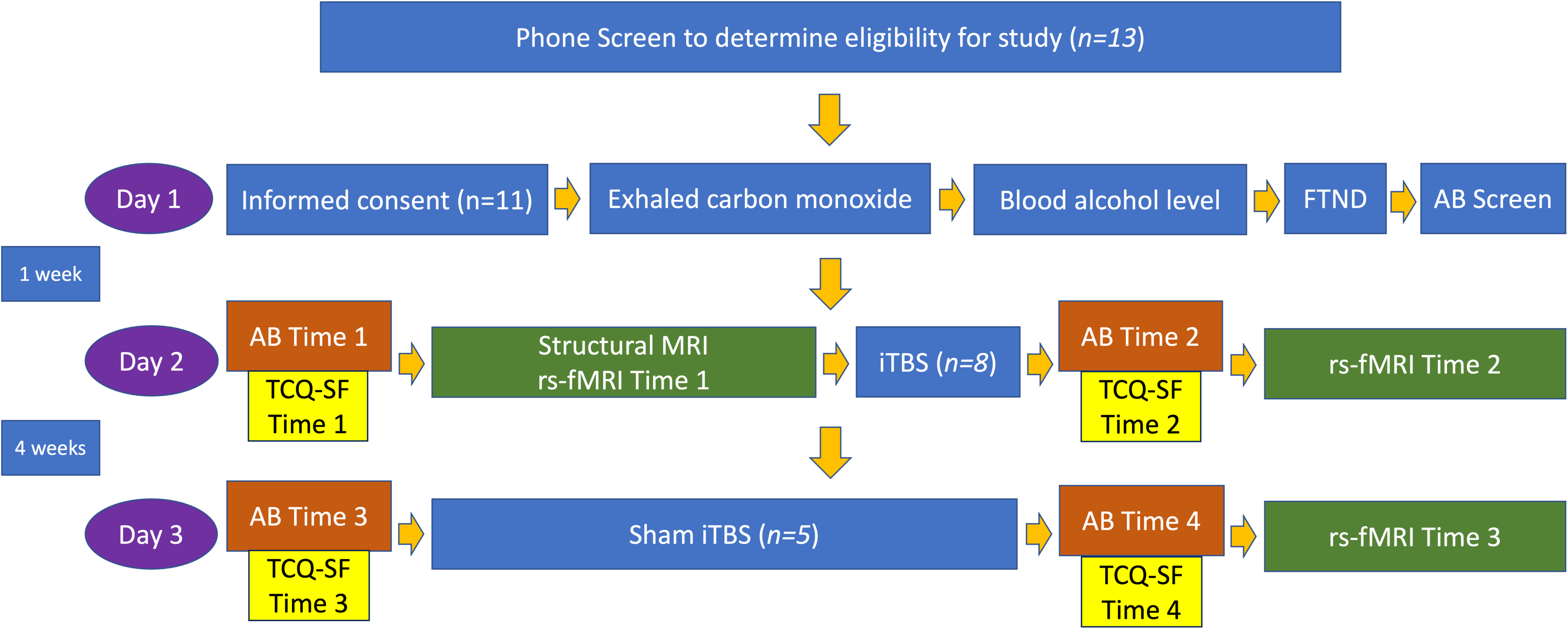
Experimental Protocol and Study Timeline. • All participants first completed a phone screen to determine preliminary study eligibility. If screened eligible via the phone screen, they were scheduled to complete an in-person assessment session (Day 1). Day 1 began with participants completing an IRB approved informed consent. Demographic information, medical history, and current and past substance use (i.e., alcohol, cannabis, methamphetamines, cocaine, opioids, and benzodiazepines) were then assessed. • Participants were instructed to abstain from smoking cigarettes or using other substances for two hours prior to start of experimental sessions on Days 1, 2 and 3 of the study. Smoking abstinence was corroborated with carbon monoxide (CO) measurement using a Smokerlyzer Breath CO Monitor (Bedfont Scientific Ltd., Rochester, England). Blood alcohol level (BAL) was also recorded using a Breathalyzer. To proceed further with study procedures on Day 1, participants were required to have a CO level less than 10 ppm and a BAL of 0. • Subsequently, they completed the Fagerstrom test for nicotine dependence (FTND) and the screener attentional bias tasks (AB Screen). If they fulfilled all inclusion and exclusion criteria and demonstrated attentional bias on the screener attentional bias task (Day 1), they were then scheduled for iTBS after a week, and sham iTBS sessions after four weeks. Participants were instructed to adhere to the same abstinence routine on both session days. • On day 2, participants performed Smokerlyzer and Breathalyzer assessments. Subsequently, they underwent structural MRI brain and resting state functional MRI brain (rs-fMRI Time 1) before completing attentional bias tasks (AB Time 1) and a cigarette craving assessment (TCQ-SF Time 1). Resting motor threshold (RMT) was then measured and iTBS was delivered at 120 % of RMT. Immediately following iTBS, participants performed attentional bias tasks (AB Time 2) along with a cigarette craving assessment (TCQ-SF Time 2). They were then transported to the MRI scanner and rs-fMRI was acquired within one hour of iTBS (rs-fMRI Time 2). • Study procedures on day 3 were same as Day 2, save for two differences: sham iTBS was delivered and rs-fMRI scans were acquired only after sham iTBS. Participants performed attentional bias tasks before and after sham iTBS sessions (AB Time 3 and AB Time 4) and rs-fMRI was acquired within one hour of sham iTBS (rs-fMRI Time 3). We assessed cigarette craving before and after sham iTBS (TCQ-SF Time 3 and TCQ-SF Time 4) respectively. Five out of eight participants returned for sham iTBS and completed Day 3 procedures.

### Attentional Bias

We used a visual probe task to measure AB, adapted for cigarette cues^56^. We used a Tobii Pro Fusion 120 Hz eye tracker (Tobii Technology, Sweden) to monitor fixation time on cigarette and neutral cues, which helped calculate cigarette cue AB. For each AB trial, images of a cigarette (C) and a matched neutral (N) cue were presented on a laptop screen, 3 cm (about 1.18 in) apart. Upon offset of the image pairs, a visual probe (X) appeared on either the left side or right side of the screen, in the same location as one of the previously presented images. Firstly, twenty trials showed images of cues encompassing people smoking cigarettes and matched neutral cues for 2000 milliseconds (ms) (about 2 seconds). Secondly, twenty trials showed images of cues encompassing cigarette paraphernalia and matched neutral cues for 2000 ms.

For each set of twenty cigarette cue AB trials, twenty filler trials consisting of twenty pairs of additional neutral cue images (N-N) were intermixed to generate a total of 40 trials. Filler trials were also presented for 2000 ms. Intervals between C-N and N-N cue pairs were 2000 ms. Average fixation time was calculated separately for each set of AB trials by summing the total fixation time for each cue type across all trials and then dividing by the total number of critical trials (i.e., 20). Cigarette cue AB for each set of AB trials was calculated by average fixation time on cigarette cues minus average fixation time on neutral cues^56^.

#### fMRI

Neuroimaging acquisition was completed with a 3T Siemens Magnetom PRISMA Scanner. Structural MRI images (MPRAGE) were acquired before iTBS. Multi-slice gradient echoplanar imaging (EPI) resting state images were acquired before iTBS (rs-fMRI Time 1), within an hour after iTBS (rs-fMRI Time 2) and within an hour after sham iTBS (rs-fMRI Time 3). Specifics of scan sequences are elucidated in Supplementary material Section 4.

#### TMS and Neuronavigation for Targeting

Structural MRI images were input into Brainsight software (Rogue Solutions, Montreal, Canada). The images were then registered to Montreal Neurological Institute (MNI) space by identifying the anterior commissure and posterior commissure in Brainsight software (Rogue Solutions, Montreal, Canada). The registered images were consequently used to generate a three-dimensional curvilinear brain model for each subject. This was used to identify the L. dlPFC (Brodmann area 46, MNI coordinate −44, 40,29) ^57,58^. During delivery of iTBS, nine minutes eleven seconds (1800 pulses) of iTBS (biphasic bursts) at 120% of resting motor threshold (RMT) was administered to the L. dlPFC using MagVenture MagPro x100 with MagOption and B65 active/placebo (A/P) coil (MagVenture A/S, Denmark). The coil position was stabilized using a holder, with guidance from Brainsight Neuronavigation (Rogue Solutions, Montreal, Canada). Sham iTBS was administered using the sham interface of the B65 A/P coil. Two stimulation electrodes (Ambu Neuroline 710) were connected to the coil and placed on the left side of the scalp to mimic the somatosensory stimulation associated with iTBS. RMT was measured prior to iTBS and sham iTBS (RMT procedure described in Section 3 of Supplementary material).

### Clinical Assessments

#### Tobacco Craving Questionnaire-Short Form (TCQ-SF)

The Tobacco Craving Questionnaire-short form (TCQ-SF) consists of 12 items rated on an 84-point visual analogue scale. Validity and reliability of the TCQ-SF is comparable to the original 47-item version ^59^. The TCQ-SF was administered before and after iTBS on Day 2 (TCQ-SF Time 1 and TCQ-SF Time 2 in **Figure 1**) and sham iTBS on Day 3 (TCQ-SF Time 3 and TCQ-SF Time 4 in **Figure 1**).

#### Fagerstrom Test for Nicotine Dependence (FTND)

The FTND is a six-item self-report measure of nicotine dependence severity. Three yes(1)/no(0) items and three multiple choice items (scored from 0 to 3) are used to calculate total severity (0-10)^60^.

### Data Analytic Strategy

#### Resting state functional connectivity (rsFC) analyses

CONN functional connectivity toolbox (version 21.a) was used for all preprocessing and neuroimaging analyses using standard procedures (see Supplementary Material Section 4). Firstly, general linear model (GLM) analyses explored if rsFC changed from two seeds (L. dlPFC and right insula) to all voxels following iTBS (rs-fMRI Time 2) versus sham iTBS (rs-fMRI Time 3) relative to baseline rsFC (rs-fMRI Time 1) in the five participants who completed all three days of the study. In first level analyses, a seed map was created for every subject using respective seeds (L. dlPFC or right insula). In second level analyses, a between conditions contrast of (0,1,-1) was used to assess rsFC within-subject differences across the three time points (rs-fMRI Time 1, rs-fMRI Time 2, rs-fMRI Time 3). All second level analyses used cluster level inferences based on Gaussian Random Field Theory ^61^. This was done using general linear models (GLM) to generate a statistical map of t-values, that were subsequently thresholded to reveal non overlapping clusters (https://web.conn-toolbox.org/fmri-methods/cluster-level-inferences). Two-sided p-values were generated using cluster-level false discovery rate (FDR) correction ^62^.

Subsequently, rsFC change from two seeds (L. dlPFC and right insula) to all voxels across from rs-fMRI Time 1 to rs-fMRI Time 2 was examined among the eight participants who only received iTBS. In first level analyses, a seed map was created for every subject using respective seeds (L. dlPFC or right insula). In second level analyses, a GLM with between conditions contrast of (1,-1) for scans (rs-fMRI Time 1, rs-fMRI Time 2) was used. In these eight participants, associations between changes in seed based rsFC following iTBS (rs-fMRI Time 1 versus rs-fMRI Time 2) and changes in fixation time on cues of people smoking cigarette (AB Time 1 versus AB Time 2) were explored. A GLM was specified with between conditions contrast of (−1,1) for rs-fMRI Time 1 and rs-fMRI Time 2, respectively, and a between subjects’ contrast of (−1,1) for fixation time on cues encompassing people smoking cigarettes acquired as part of AB Session 1 and AB Session 2 respectively.

#### Attentional Bias Analyses

Cigarette cue AB was calculated separately for each set of AB trials, by subtracting the average fixation time on neutral cues from the average fixation time on cigarette cues. Fixation time on cigarette cues for each session and resultant cigarette cue attentional bias score were entered into separate linear fixed effects regression models. Dependent variables for these models were fixation time on cigarette cue and cigarette cue AB, respectively. Fixed effects for both models were type of cigarette cues (people smoking cigarettes versus cigarette paraphernalia), time (before or iTBS/sham iTBS), intervention arm (iTBS versus sham iTBS) and FTND scores. Subject ID was a grouping variable in both models. Further details on regression models can be found in Supplementary material section 5.

#### Cigarette craving analyses

TCQ-SF scores were entered into a linear fixed effects regression model. Outcome variables for this model was TCQ-SF score. Predictor variables for this model were time (before or after iTBS/sham iTBS), intervention arm (iTBS versus sham iTBS), and FTND scores. Further details on regression models can be found in Supplementary material section 5.

## Results

### Seed-Based Resting State Functional Connectivity

General linear models (GLMs) comparing differences in rsFC between iTBS (rs-fMRI Time 2) and sham iTBS (rs-fMRI Time 3), relative to rs-fMRI Time 1, suggested that rsFC significantly increased between the L. dlPFC and a cluster comprising bilateral medial prefrontal cortex (MPFC), bilateral temporal poles, and pons [*t*(4)=28.87, *p*=0.0007] (**Figure 2 and Table 2**) and significantly increased between the right insula and a cluster comprising the right occipital pole and inferior division of right lateral occipital cortex [*t*(4)=15.40, *p*=0.04] (**Figure 2 and Table 2**). GLMs comparing change in rsFC from pre-to-post iTBS (rs-fMRI Time 1 and Time 2), suggested that rsFC significantly increased between the L. dlPFC and a cluster comprising the right frontal pole and right paracingulate gyrus [*t*(7)=10.75, *p*=0.02] (**Figure 2 and Table 2**) and significantly increased between the right insula and in two voxel clusters – one comprising the right precentral gyrus and right postcentral gyrus, [*t*(7)=9.34, *p*=0.04] and one comprising the left precentral gyrus and left postcentral gyrus [*t*(7)=11.18, *p*=0.04] (**Figure 2 and Table 2**).

**Figure 2.**
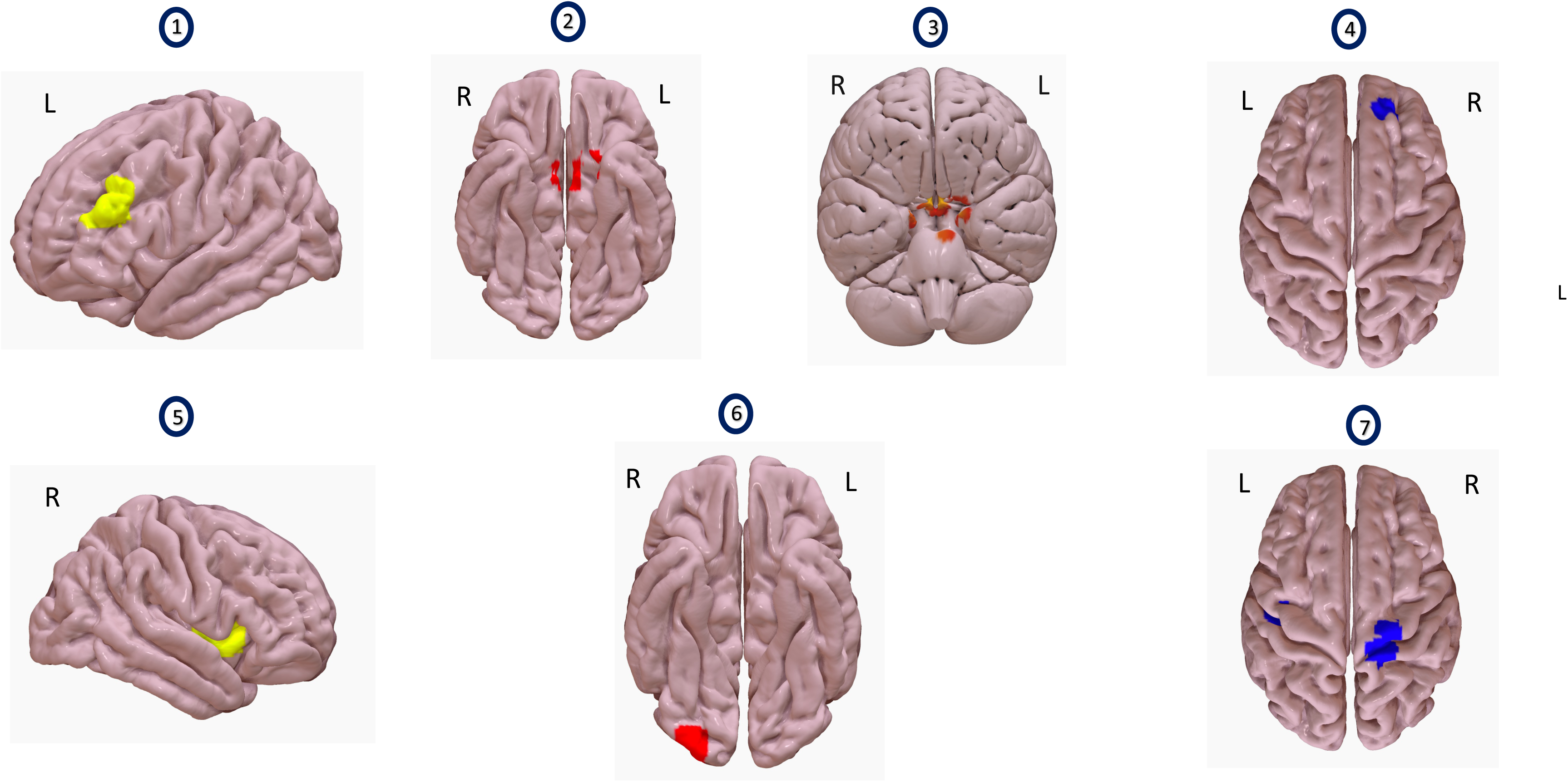
Comparing resting state functional connectivity (rsFC) changes between iTBS and sham iTBS. • Images 1 and 5 show seeds used for analyses. Image 1 shows the left dlPFC and image 5 shows the right insula. • Images 2 and 3 show a voxel cluster where resting state functional connectivity (rsFC) with L. dlPFC as a seed increased, with iTBS compared to sham iTBS (n=5). This cluster (red) encompasses bilateral medial prefrontal cortex (Image 2), bilateral temporal poles and pons (Image 3). • Image 4 shows a voxel cluster where rsFC with L.dlPFC as a seed increased, after receiving iTBS, compared to before (n=8). The voxel cluster (blue) is situated over right frontal pole and right paracingulate gyrus. • Image 6 shows a voxel cluster where rsFC with right insula as a seed increased, after receiving iTBS compared to sham iTBS (n=5). The voxel cluster (red) is situated in the right occipital pole and right lateral occipital cortex. • Image 7 shows a voxel cluster where rsFC with right insula as a seed changed, after receiving iTBS, compared to before (n=8). The figure shows a voxel cluster (blue) situated over bilateral precentral and bilateral postcentral gyri.

**Table 2.** Resting State Functional Connectivity (rsFC) Changes with iTBS/Sham iTBS (Two seeds - left dorsolateral prefrontal cortex or L. dlPFC and right insular cortex to all voxels)

| Seed Region | Between Conditions Contrast Applied | Voxel Number | MNI coordinates | *T* Statistic | *Beta* * |
| --- | --- | --- | --- | --- | --- |
| Changes in seed -voxel rsFC between rs-fMRI Time 2 and rs-fMRI Time 3 (*n*=5) |  |  |  |  |  |
| L. dIPFC | GLM (rs-fMRI Time 1, rs-fMRI Time 2, rs-fMRI Time 3; 0, 1, -1) (*n*=5) | 96 | (+6, -2, -24) bilateral medial prefrontal cortex, bilateral temporal poles, and pons. | 28.87 | 0.18 |
| R. Insula | GLM (rs-fMRI Time 1, rs-fMRI Time 2, rs-fMRI Time 3; 0, 1, -1) (*n*=5) | 49 | (+30, -92, -10) right occipital pole and inferior division of right lateral occipital cortex | 15.40 | 0.29 |
| Changes in seed -voxel rsFC between rs-fMRI Time 1 and rs-fMRI Time 2 (*n*=8) |  |  |  |  |  |
| L. dIPFC | GLM (rs-fMRI Time 1, rs-fMRI Time 2; -1,1 (*n*=8)) | 79 | (+14, +50, +22) right frontal pole and right paracingulate gyrus | 10.75 | 0.21 |
| R. Insula | GLM (rs-fMRI Time 1, rs-fMRI Time 2; -1,1 (*n*=8)) | 64 | (+24, -20, +66) right precentral gyrus and right postcentral gyrus | 9.34 | 0.15 |
|  |  | 61 | (-40, -14, +34) left precentral gyrus and left postcentral gyrus | 11.18 | 0.15 |
| L. dIPFC | GLM (rs-fMRI Time 1, rs-fMRI Time 2;-1,1; ** Fixation on Cigarette Cues AB Time 1, Fixation on Cigarette Cues AB Time 2; -1,1), (*n*=8) | No significant results |  |  |  |
| R. Insula | GLM (rs-fMRI Time 1, rs-fMRI Time 2; -1,1; ** Fixation on Cigarette Cues AB Time 1, Fixation on Cigarette Cues AB Time 2; -1,1), (n=8) | 181 | (-38, -38, +48) right superior parietal lobule, right postcentral gyrus, right anterior division of supramarginal gyrus. | -8.07 | -0.0023 |
|  |  | 79 | (+26, -36, +46) left superior parietal lobule, left postcentral gyrus, left anterior division of supramarginal gyrus, left posterior division of supramarginal gyrus. | -12.37 | -0.0018 |

rsFC – resting state functional connectivity
GLM – general linear model
All results thresholded at *false discovery rate (FDR)* corrected cluster threshold *p value* <0.05 and uncorrected voxel threshold *p value* <0.0001.
\*- *Beta* represents the regression coefficient displaying the average difference in connectivity between two or three conditions (measured with Fisher-transformed correlation coefficients), depending on the comparison being done. Connectivity measured is between the seed (L. dlPFC or right insula) and respective significant voxel cluster in each row.
\*\* - Between-subjects' contrast

### Attentional Bias and Self-reported Craving

Linear fixed effects models revealed that, across both attention bias cigarette cues, the severity of nicotine dependence (FTND) was significantly associated with changes in fixation time on cigarette cues with iTBS and sham iTBS [*t (*43) = 2.27, *p*=0.03] (**Supplementary material Table 1**). FTND was also significantly associated with changes cigarette cue AB with iTBS and sham iTBS [*t (*43) = 17.14, *p*=0.01] (**Supplementary material Table 2**). Severity of cigarette cravings (TCQ-SF) was not significantly associated with fixation time on cigarette cues or cigarette cue AB (**Supplementary material Table 3**).

Paired t-tests showed that fixation time and attentional bias to cues of people smoking cigarettes significantly reduced following iTBS, [*t (*15) =4.20, *p*=0.04] and [*t (*15) =3.14, p=0.02], respectively. See **Table 3 and Figure 3**. Similarly, nicotine craving scores reduced from a mean of 55.88 (*SEM* 3.95) to 43.13 (*SEM* 4.68) with iTBS [*t (*15) =3.02, *p*=0.02]. With sham iTBS it reduced from a mean of 53.2 (*SEM* 4.77) to 48.40 (*SEM* 6.84) [*t (*9) =1.24, *p*=0.28].

**Figure 3.**
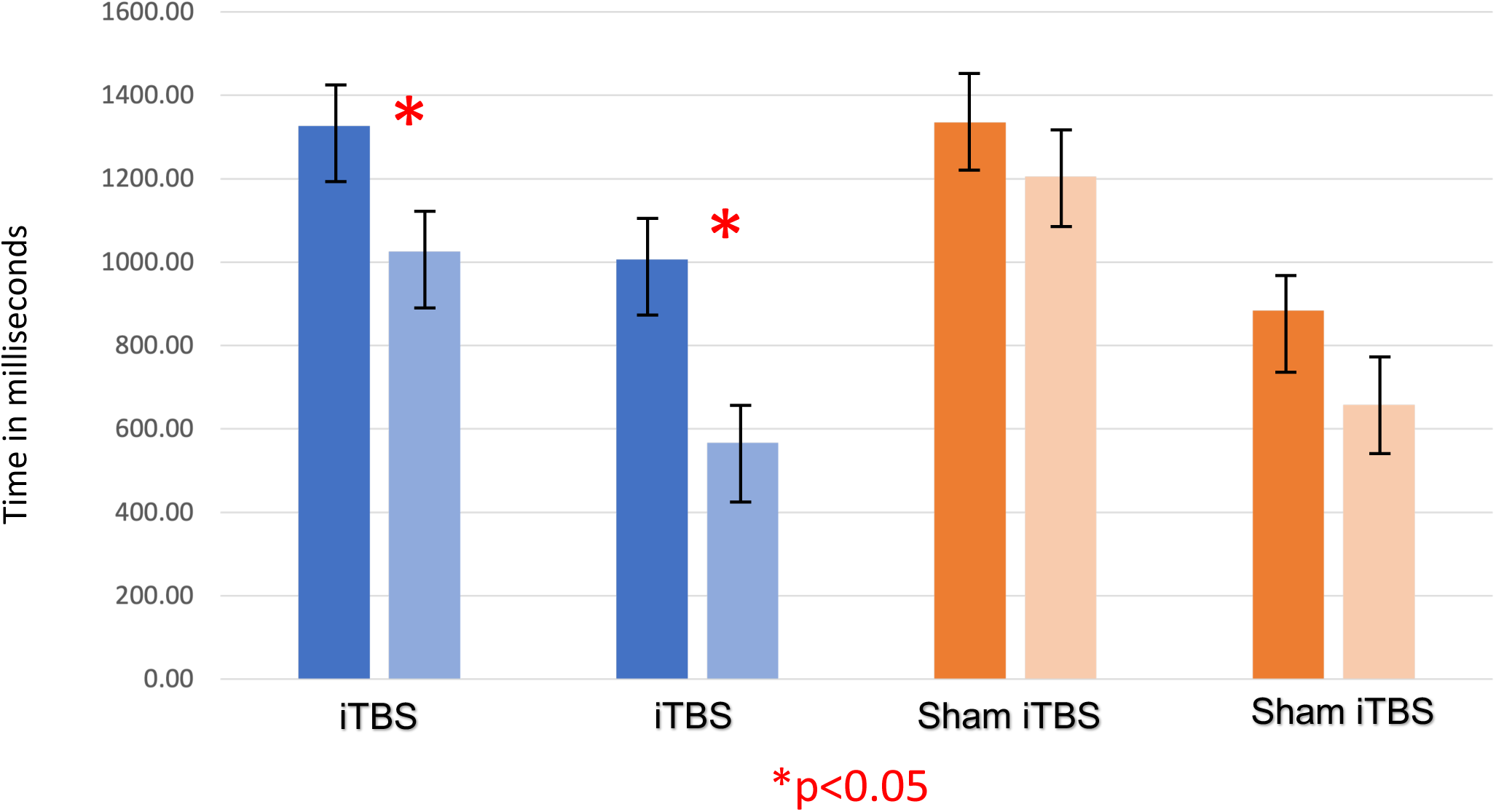
Changes in Fixation Time and Cigarette Cue Attentional Bias (AB) with iTBS/Sham iTBS. • The first two bars (shades of blue) represent gaze fixation on cues encompassing people smoking cigarettes, acquired before and after iTBS (AB Time 1 and AB Time 2). • The third and fourth bars (shades of blue) represent attentional bias for on cues encompassing people smoking cigarettes (measured by subtracting gaze fixation on neutral cues from gaze fixation on smoking cues) acquired before and after iTBS. • The fifth and sixth bars (shades of orange) represent gaze fixation on the same cues, acquired before and after sham iTBS (AB Time 3 and AB Time 4). • The seventh and eighth bars represent attentional bias for on cues encompassing people smoking cigarettes acquired before and after sham iTBS. • As observed, iTBS showed a significant decrease in gaze fixation on on cues encompassing people smoking cigarettes and consequently on attentional bias also, which was not seen with sham iTBS.

**Table 3.** Ad Hoc T-tests to Compare Changes in Fixation Time and Attentional Bias with iTBS/Sham iTBS.

| AB Session | Cues Encompassing People Smoking Cigarettes |  |  | Cigarette Paraphernalia Cues |  |  |
| --- | --- | --- | --- | --- | --- | --- |
|  | Fixation Time on Cigarette Cues (Milliseconds)<br>Mean (SEM) | Fixation Time on Neutral Cues (Milliseconds)<br>Mean (SEM) | Attentional Bias (Milliseconds)<br>Mean (SEM) | Fixation Time on Cigarette Cues (Milliseconds)<br>Mean (SEM) | Fixation Time on Neutral Cues (Milliseconds)<br>Mean (SEM) | Attentional Bias (Milliseconds)<br>Mean (SEM) |
| iTBS |  |  |  |  |  |  |
| AB Time 1 | 1326.93<br>(105.77) | 319.88 (58.06) | 1007.05<br>(148.70) | 1110.33 (88.71) | 439.91 (55.40) | 670.43<br>(139.26) |
| AB Time 2 | 1026.05<br>(100.90) | 459.16 (50.98) | 566.89<br>(121.70) | 919.14 (96.65) | 476.05 (54.48) | 443.10<br>(136.37) |
| *p Value* | 0.004 * | 0.097 | 0.016 * | 0.128 | 0.671 | 0.266 |
| *Effect Size (Cohen's d) \*\** | 2.17 |  | 1.6 |  |  |  |
| Sham iTBS |  |  |  |  |  |  |
| AB Time 3 | 1334.98 (59.10) | 450.38 (54.17) | 884.60<br>(112.77) | 1025.80 (88.33) | 497.42 (54.58) | 528.38<br>(139.39) |
| AB Time 4 | 1205.17 (73.06) | 546.50 (84.03) | 658.67<br>(148.63) | 845.88 (101.54) | 538.32 (80) | 307.56<br>(135.26) |
| *p Value* | 0.191 | 0.293 | 0.377 | 0.164 | 0.373 | 0.061 |

Presented values in cells are the mean values in milliseconds. Within parentheses in every cell is the standard error of the mean (SEM).
\**p value* ≤ 0.05.
\*\* *Cohen's d* was calculated only for comparisons with significant *p* values.

### Correlation Between Fixation Time on Cigarette Cues and Seed-Based rsFC

A GLM suggested that decrease in fixation time to cues of people smoking cigarettes was significantly associated with increases in rsFC between the right insula and two clusters comprising right postcentral gyrus [*t (*7) = −8.07, *p*=0.000005] and left postcentral gyrus [*t (*7) = −12.37, *p*=0.005] respectively following iTBS (rs-fMRI 1 and rs-fMRI 2) (**Table 2 and Figure 4**).

**Figure 4.**
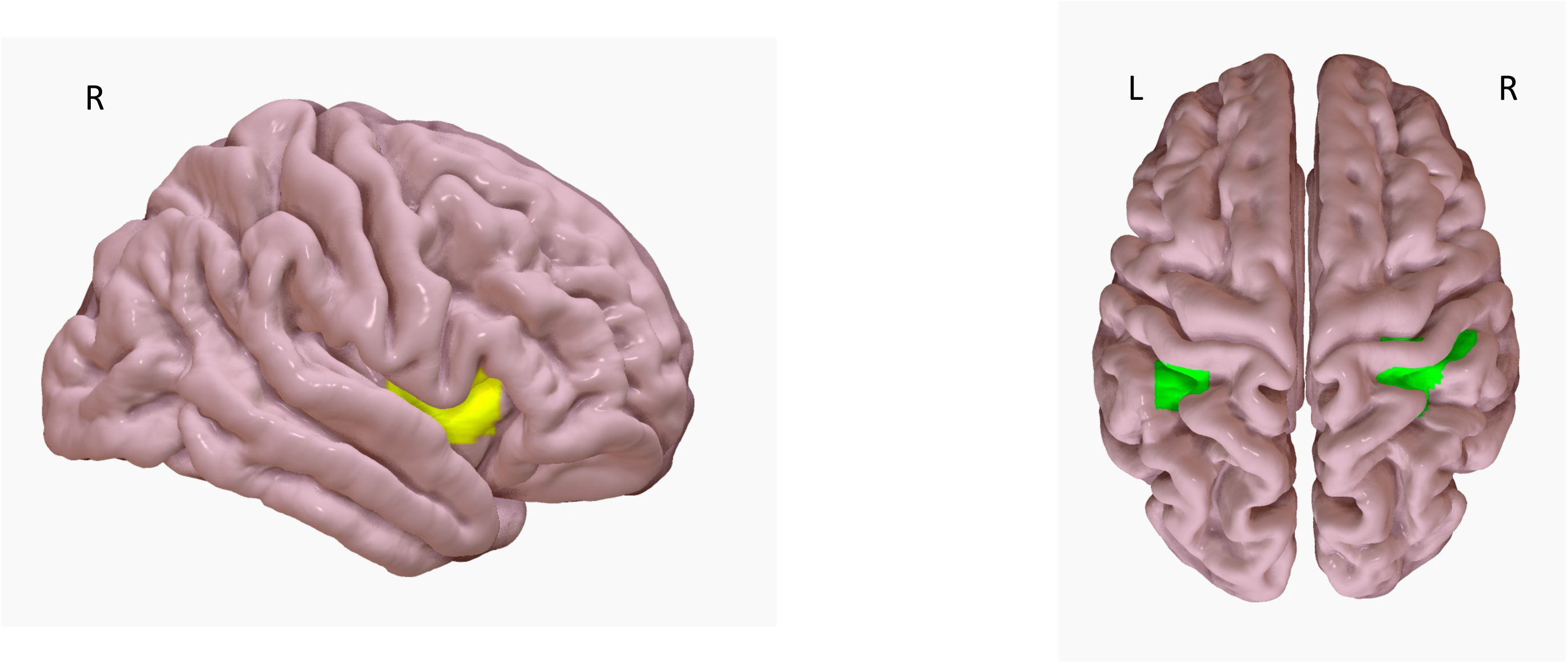
Correlation Between Fixation Time on Cues of People Smoking Cigarettes and rsFC. • Figure 4 shows two voxel clusters which had significant rsFC reduction with right insula as a seed (yellow) with reduction in gaze fixation time with iTBS. • The voxel clusters encompass bilateral superior parietal lobules and supramarginal gyri (green).

## Discussion

This is the first study to examine the effects of iTBS on rsFC, cigarette craving, and cigarette cue attentional bias in PLWHA who smoke cigarettes. We found that, compared to sham iTBS, rsFC between the L. dlPFC and a voxel cluster comprising the mPFC and pons increased more following iTBS of the L. dlPFC. We also found that rsFC between the right insula and right occipital cortex increased more following iTBS compared to sham iTBS.

With iTBS, we also saw increased rsFC between the L. dlPFC and the the right frontal pole, right paracingulate gyrus, as well as between the right insula and bilateral precentral and postcentral gyri (somatosensory cortices). Cigarette craving and cigarette cue attentional bias significantly reduced following iTBS but not following sham iTBS, though the magnitude of these changes was not significantly different across the two experimental groups. Lastly, reductions in cigarette cue fixation time following iTBS were associated with increases in rsFC between the right insula and the right and left superior parietal lobules, the right and left postcentral gyri (somatosensory cortices) and the right and left supramarginal gyri.

Our results extend literature on functional activation changes following TMS of the L.dlPFC in people who smoke. Two previous studies reported that TMS of the L.dlPFC in smokers reduced BOLD activity in the right medial orbitofrontal cortex (OFC), ^45^ and right insula^44^. The present findings suggest that rsFC of the mPFC, which included the medial OFC, and right insula may also be effected by TMS of the L.dlPFC; though the present study utilized iTBS instead of high frequency TMS. Consistent with prior TMS research ^41–43^, the present iTBS findings and past TMS research with people who smoke suggest that brain stimulation can modulate functional connectivity and functional activation of regions and networks distal from the site of stimulation.

Although measurement of cigarette cue attentional bias provided a behavioral measure to compare effects of iTBS versus sham iTBS, it is possible that this facet may have unintentionally increased craving secondary to cue presentation. Since cigarette cue attentional bias was measured immediately before and after iTBS/sham iTBS session, it is possible that cigarette cue presentation may have influenced the brain state at the time of stimulation ^63,64^. In this context, the finding that rsFC between the right insula and right lateral occipital cortex increased after iTBS is consistent with cigarette cue reactivity task studies, which reported that functional activation of the occipital cortex was positively associated with reactivity towards smoking cues in people who smoke cigarettes ^54,65^. Increased rsFC between L. dlPFC and right frontal pole and right paracingulate gyrus following iTBS extends results from a meta-analysis examining neural correlates of cigarette cue reactivity. This meta-analysis showed the left anterior cingulate gyrus, paracingulate gyrus and dorsal cingulate cortex to be instrumental when examining fMRI correlates of reactivity towards smoking cues in people who smoke^66^.

Our regression models suggested that changes in fixation time (**Supplementary material table 1**), cigarette cue attentional bias (**Supplementary material table 2**), and cigarette craving (**Supplementary material table 3**) were not significantly different between iTBS and sham iTBS. Nonetheless, we showed a significant reduction in craving, cigarette cue fixation time and attentional bias following iTBS and not following sham iTBS. The decrease in cigarette craving has been shown in previous TMS studies^16^ and one iTBS study^29^. However, our study is the first to show decrease in cigarette cue attentional bias with iTBS.

The decrease in cigarette cue fixation time to cues of people smoking cigarettes following iTBS was also significantly associated with increases in rsFC between the right insula and clusters comprising the right and left somatosensory cortices in addition to the right and left posterior parietal cortices. This extends the literature on the role of right insula and parietal cortex in craving. Structural white matter connectivity between posterior parietal cortex and right insula can modulate craving^74^. Right anterior insula is crucial for interceptive awareness and salience^67,68^. The posterior parietal cortices modulate selective attention, in addition to the somatosensory cortices being important for perceptual awareness^69,70^. When viewing cigarette and neutral cues for cigarette cue attentional bias measurement, the lateral posterior parietal cortex in conjunction with the right anterior insula integrates perceptual awareness of cues with internal physiological awareness, consequently modulating sustained attention^71,72^. These facets become crucial to craving and attentional bias as well^73^.

We delivered 1800 pulses of iTBS to L.dlPFC, based on two seminal studies in major depressive disorder (MDD) which used the same dosing strategy in an accelerated fashion and showed a remission rate of approximately 90% in treatment resistant MDD^32,33^. We used a stimulus intensity of 120% of RMT based on a previous iTBS trial^31^. Prior to this study, neither of these TBS dosing parameters have been applied to TMS studies in people who smoke cigarettes (regardless of HIV status). Moreover, the present study was the first to use neuronavigational targeting with structural MRI in PLWHA who smoke cigarettes.

We delivered actual and sham iTBS in a single blind fashion. Although patient expectations and placebo effects could have influenced results, we separated active and sham iTBS sessions by four weeks, which should have limited carryover effects ^75^. Nonetheless, replication and extension efforts should include double blinding. In an ideal design, we would have acquired a resting state fMRI scan before sham iTBS too. This is a limitation of our study. Although we measured cigarette cue attentional bias immediately after iTBS/sham iTBS, there was an average of an hour’s delay before they got to the scanner, and this may have influenced our results as well.

Future research using online (with cue presentation) vs. offline (at rest without prior provocation) iTBS could explore the potential moderating effects of cue presentation on iTBS in people who smoke cigarettes.

## Conclusions

iTBS to the L. dlPFC in PLWHA who smoke increased rsFC between L.dlPFC and bilateral medial prefrontal cortex and pons, and increased rsFC between right insula and right occipital cortex. Despite a small sample size, iTBS decreased cigarette cue fixation time, cigarette cue attentional bias and cigarette craving. We also showed a negative association between change in cigarette cue fixation time to cues of people smoking cigarettes and rsFC between the right insula and clusters comprising the right and left somatosensory cortices in addition to the right and left posterior parietal cortices. Our results warrant a powered prospective trial evaluating iTBS as a therapeutic intervention for smoking cessation, along with an optimal rsFC biomarker to guide treatment response.

## Data Availability

All data produced in the present study are available upon reasonable request to the authors

## Acknowledgment

This work was supported by the National Institutes of Health (grant numbers AA026255, TR001997, CA225419, MH129832, CX00293, MH111977) and University of Kentucky College of Medicine.

