## Supplementary Material for "Intermittent Theta Burst Stimulation (iTBS) and Resting State Functional Connectivity in People Living with HIV/AIDS (PLWHA) Who Smoke Tobacco Cigarettes"

Section 1. Recruitment. We screened 13 potential participants and consented eleven. Two participants were excluded during screening due to a history of seizures. Two participants were excluded after consenting because they did not demonstrate attentional bias for cigarette cues during initial assessment (AB Screen in Figure 1). One dropped out of the study after Day 1 due to personal reasons.

Section 2. Inclusion criteria- 1) Patients enrolled in the Bluegrass Clinic; 2) 18-60 years of age; 3) male or female gender 4) Able to read, understand and communicate in English; 5) willing and able to abstain from drug use other than Suboxone; 6) exhaled CO < 10 ppm on study day; 7) Stabilized on maintenance buprenorphine if having comorbid opioid use disorder.

Exclusion criteria. Positive pregnancy test for females, traumatic brain injury, history of seizure disorder, history of or current diagnosis of psychosis, intracranial metal shrapnel, previous adverse effect with TMS, sub-threshold consistency while performing behavioral tasks, failure to show baseline attentional bias to smoking versus neutral cues.

Section 3.Resting motor threshold (RMT). We used the B60 coil (figure of eight) to measure motor threshold. Resting motor threshold (RMT), is defined as minimum single pulse intensity required to produce a motor evoked potential (MEP) greater than or equal to 50 mV on more than five out of 10 trials from the contralateral first dorsal interosseus muscle (FDI) while the subject is resting. TMS pulses of 1 Hz were administered in a consecutive manner, using increments of 5% of stimulator output starting from 35%, until we were able to consistently elicit a motor evoked potential (MEP) of 50 microvolts from the right FDI. This was done using Brainsight 2 EMG pod (Rogue Research, Montreal, Canada), Brainsight EMG amplifier (Rogue Research, Montreal, Canada) and three foam electrodes (Kendall) applied to consistent locations to measure activity from right FDI. The location on the scalp that produced consistent responses as detailed above was called the ‘motor hotspot.’ The location of the motor hotspot was recorded in Brainsight neuronavigation with the help of a software interface. Subsequently the same coil and 1 Hz pulses were used in conjunction with the PEST algorithm to identify a precise resting motor threshold. We did not measure the scalp to cortex distance and utilized a stimulation threshold of 120% to compensate for any effects this may have had on our outcomes. This threshold was also used in the THREE-D trial that compared iTBS to 10Hz TMS.

Section 4. Scan sequences and preprocessing. We acquired structural MRI brain using magnetization-prepared rapid gradient-echo (MPRAGE) sequence with the following parameters: - 208 slices, FoV 256 mm X 240 mm, slice thickness 0.8 X 0.8 X 0.8 mm3, TR 2500 milliseconds, TE 2.22 milliseconds, flip angle 8 degrees. The sequence lasted 6.54 minutes. Multi-slice gradient echo planar imaging (EPI) sequence was used for all resting state (rs-fMRI) scans with the following parameters - 72 slices, FoV 208 mm X 208 mm, slice thickness 2 X 2 X 2 mm3, TR 800 milliseconds, TE 37 milliseconds, flip angle 52 degrees, 488 volumes, run time 6.40 minutes. MRI preprocessing and analyses – All structural brain MRIs and resting state fMRI brain scans were processed and analyzed using CONN functional connectivity toolbox (version 21.a) ([www.nitrc.org/projects/conn](http://www.nitrc.org/projects/conn)), implemented in MATLAB (version R2021a; MathWorks, Inc.). As part of preprocessing, the functional images were 1) realigned, 2) unwarped, 3) corrected for motion and slice timing, 4) examined for functional outliers, 5) segmented, and normalized to Montreal Neurological Institute (MNI) space, 6) resampled at 2-mm^3^, 7) and smoothed with a Gaussian kernel of 8-mm full-width at half-maximum.

A denoising step was then applied to remove the potential confounders associated with realignment and scrubbing covariates, as well as BOLD signals from white matter and cerebrospinal fluid (CSF) masks. The band-pass filtering between 0.008 Hz and 0.09 Hz (the default setting of CONN toolbox) was employed to reduce low-frequency drift and high-frequency respiratory and cardiac noise. Regions of interest (ROIs) for analyses were chosen from the Harvard-Oxford Atlas comprising 48 cortical and 21 subcortical ROIs (total of 132 ROIs). An extra ROI for the left dlPFC was constructed in Conn Toolbox based on site of stimulation (MNI coordinate –44,29,40) and a sphere radius of 10 mm.

Seed to voxel connectivity analyses was performed using left dlPFC seed (constructed ROI) and right insula (HOA atlas ROI) as seed ROIs. Bivariate correlations and canonical hemodynamic response function (HRF) weighting were used.

Section 5. Linear mixed effects models

Supplemental Table 1 – Regression Model with Fixation Time on Smoking Cues as Outcome Variable [*R^2^* (43) = 0.54].

Model – Fixation time on cigarette cues (milliseconds) ~ 1 + attentional bias (AB) task (one task measuring bias for people smoking cigarette cues versus neutral cues and another task measuring bias for cigarette paraphernalia cues versus neutral cues) + time (before or after iTBS/sham iTBS) + intervention arm (active iTBS versus sham iTBS) + Fagerstrom test for nicotine dependence (FTND) scores + AB task * time + AB task * intervention arm + AB task * time + time * intervention arm + 1|Subject ID.

| Name | *Estimate (Beta)* | *Standard Error* | *T Statistic* | *Degrees of freedom* | *p value* |
| --- | --- | --- | --- | --- | --- |
| Intercept | 2039.7 | 780.57 | 2.61 | 43 | 0.01 |
| Task (People/Objects) | -393.48 | 487.82 | -0.81 | 43 | 0.42 |
| Time (Pre/post) | -741.41 | 487.82 | -1.52 | 43 | 0.14 |
| Arm (Active/sham) | -348.46 | 526.05 | -0.66 | 43 | 0.51 |
| FTND Scores | 45.71 | 20.179 | 2.27 | 43 | 0.03 |
| Task* Time | 269.47 | 308.52 | 0.87 | 43 | 0.39 |
| Task* Arm | 67.197 | 332.39 | 0.20 | 43 | 0.84 |
| Time*Arm | 330.85 | 332.39 | 0.99 | 43 | 0.33 |
| Task*Time*Arm | -159.79 | 210.22 | -0.76 | 43 | 0.45 |

Supplemental Table 2 – Regression Model with Cigarette Cue Attentional Bias as Outcome Variable [*R^2^* (43) = 0.23].

Model -Cigarette Cue Attentional bias (milliseconds) ~ 1 + type of attentional bias (AB) task performed (one task measuring bias for people smoking cigarette cues versus neutral cues and another task measuring bias for cigarette paraphernalia cues versus neutral cues) + time (before or after iTBS/sham iTBS) + intervention arm (active iTBS versus sham iTBS) + Fagerstrom test for nicotine dependence (FTND) scores + AB task * time + AB task * intervention arm + AB task * time + time * intervention arm + 1|Subject ID.

| Name | *Estimate (Beta)* | *Standard Error* | *T Statistic* | *Degrees of freedom* | *p value* |
| --- | --- | --- | --- | --- | --- |
| Intercept | 2329.2 | 1394.5 | 1.67 | 43 | 0.10 |
| Task (People/Objects) | -737.57 | 880.74 | -0.84 | 43 | 0.41 |
| Time (Pre/post) | -1074.9 | 880.74 | -1.22 | 43 | 0.23 |
| Arm (Active/sham) | -589.62 | 949.2 | -0.62 | 43 | 0.54 |
| FTND Scores | 44.704 | 17.14 | 2.61 | 43 | 0.01 |
| Task* Time | 420.54 | 557.03 | 0.75 | 43 | 0.45 |
| Task* Arm | 188.12 | 600.12 | 0.31 | 43 | 0.76 |
| Time*Arm | 421.94 | 600.12 | 0.70 | 43 | 0.49 |
| Task*Time*Arm | -207.72 | 379.55 | -0.55 | 43 | 0.59 |

Supplemental Table 3 – Regression Model with Cigarette Craving as Outcome Variable [*R^2^* (43) = 0.57].

Model- Craving ~ 1 + time (before or after iTBS/sham iTBS) + intervention arm (active iTBS versus sham iTBS) + Fagerstrom test for nicotine dependence (FTND) scores + time * intervention arm + 1|Subject ID.

| Name | *Estimate (Beta)* | *Standard Error* | *T Statistic* | *Degrees of freedom* | *p value* |
| --- | --- | --- | --- | --- | --- |
| Intercept | 68.53 | 18.41 | 3.72 | 19 | 0.001 |
| Time (Pre/post) | -21.11 | 11.20 | -1.89 | 19 | 0.07 |
| Arm (Active/sham) | -14.76 | 11.40 | -1.30 | 19 | 0.21 |
| FTND | 2.62 | 1.15 | 2.28 | 19 | 0.03 |
| Time*Arm | 8.15 | 7.38 | 1.10 | 19 | 0.28 |
